# REPEATABILITY OF A CALIBRATED DIGITAL SPECTROPHOTOMETER FOR DENTAL SHADE EVALUATION IN CURRENT, FORMER AND NEVER SMOKERS – STUDY PROTOCOL

**DOI:** 10.1101/2021.01.16.21249901

**Authors:** Gianluca Conte, Sebastiano Antonio Pacino, Salvatore Urso, Rosalia Emma, Fabio Cibella, Eugenio Pedullà, Riccardo Polosa

**Author notes:** **Corresponding author:** Dr. Gianluca Conte, Department of General Surgery and Medical-Surgical Specialties, University of Catania, Catania, Italy.

## Abstract

Despite the negative impact of cigarette smoking on oral health and teeth appearance, there is no data available on dental shade changes in smokers who quit smoking. Dental discoloration caused by smoking may be permanent, with minimal restoration after stopping smoking. If this is valid, former smokers can show dental shade values equivalent to those of current smokers.

The aim of this study is to compare the dental shade assessment by digital spectrophotometry (VITA Easyshade V) in current, former and never smokers and to verify the short (7 days) and long-term (30 days) repeatability of these measurements.

Confirmation of good reproducibility of VITA Easyshade V with clear objective discrimination of dental shade measurements among current, former, and never smokers will improve the power of this measurement giving more confidence in clinical research findings of dental shades in these populations.

It is also anticipated that results from the study will expand the application of this measurements to include medical and regulatory research applied to combustion-free tobacco products (e.g. e-cigarettes, heated tobacco products, oral tobacco/nicotine products, etc.), smoking cessation medications, and to consumer care product for oral hygiene and dental aesthetics.

## Introduction

Teeth color and appearance are affected by several factors, including lighting conditions, translucency, opacity, light scattering, gloss, and human eye perception (1). Indeed, differences in tooth color exist among people, among teeth in the same person and even within the same tooth. Visual color determination of a natural tooth can vary substantially thus leading to unreliable color assessment (2,3). For example, the nature of the surrounding light in the room and its type, power and input angle is known to be an important factor influencing color perception (4).

Tooth discoloration is generally classified as intrinsic or extrinsic, based on its etiology. Intrinsic tooth discolorations are associated with developmental abnormalities (such as fluorosis, enamel hypoplasia), whereas extrinsic discolorations are often caused by colorants from dietary (such as red wine, coffee, tea) and environmental sources (5).

Smoking tobacco is an important risk factor for poor oral health and discoloration of the tooth (6). It is well known that pigments that can stain and discolor human tissues, including teeth, skin and fingernails, are found in the particulate matter of tobacco combustion, commonly known as nicotine-free dry particulate matter or tar (7,8,9). The intensity and degree of smoke-related discoloration of the teeth may depend on the length and frequency of consumption of tobacco cigarettes (10) as well as on the intrinsic characteristics of the tooth (such as the presence of rough/irregular enamel surfaces) that can promote tar adhesion to dental surfaces (11).

Visual determination of natural teeth’s color tone is inaccurate and tooth shade guides have been introduced to minimize variability. Tooth shade guides are traditionally used by dentist to assess teeth colour and to determine how much whiter teeth can become after using various interventions (e.g. different toothbrush, toothpaste, mouthwash or home whitening solutions). However, studies with tooth shades guide have shown very poor interobserver correspondence (12,13) with accuracy for human visual determination of teeth shades faring as low as 25-35% (14,15). Clearly, human shade assessment cannot be considered for high quality research and more accurate and more reproducible methods are necessary.

In the last decade, innovative color measurement technologies have been introduced to allow more consistency of dental shade measurement. In particular, spectrophotometers are considered as the most accurate, useful and flexible device for colour matching (16). Paul et al (15) reported that spectrophotometers offered an average 33% increase in accuracy and an overall more accurate dental shade match in 93.3% of cases compared to observation performed by human eye. Their accuracy is a consequence of technological improvements, primarily consisting in the introduction of an internal light source allowing for contact measurements that avoid subjective variables of color analyses (17) and the influence of external source of light (18).

The majority of the studies on tooth color determinations with digital devices are based on CIE L*a*b* color parameters, established in 1976 by the Commission Internationale de l’Eclairage (CIE) (19). The L*a*b* color space is described as: Lightness (L*) is a measure of the lightness of an object, which represents the quantity of light reflected by an object and ranges from 0 (black) to 100 (white); parameter a* is a chromaticity measure of greenness/a < 0/ or redness/a > 0/; parameter b* is a chromaticity measure of blueness /b < 0/ or yellowness /b> 0/. In this color space, the color difference between 2 objects with colors C_1_= (L*_1_, a*_1_, b*_1_) and C_2_= (L*_2_, a*_2_, b*_2_) is defined

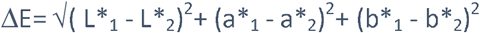

According to Johnston et Al (20) the acceptance limit for tooth colour matching in oral environment is 3.7 ΔE units, beyond this limit differences could be considered as clinically perceivable.

No data is available about changes in dental shade in smokers who quit smoking. The impairment of dental discolorations caused by smoking may be permanent, with limited restoration after stopping smoking. If this is true, former smokers should show dental shade values similar to those of current smokers.

The objective of the study is to compare the dental shade assessment by Vita Easyshade V (VITA Zahnfabrik, Bad Säckingen, Germany) in current, former and never smokers and to verify the short (7 days) and long-term (30 days) repeatability of these measurements.

## METHODS

### Study Population

The study population will consist of three study groups identified among a pool of subjects who attended a smoking cessation clinic (CPCT, Centro per la Prevenzione e Cura del Tabagismo of the University of Catania) in the previous 2 yrs or contacted among hospital staff.

Study group 1 consisted of current smokers, defined as smokers of > 10 cigarettes per day with an exhaled carbon monoxide (eCO) level of ≥ 7 ppm.

Study group 2 consisted of former smokers, defined as quitters of at least 12 months and who were still abstinent when contacted for enrollment, with an eCO level of < 7 ppm.

Study group 3 consisted of never smokers, defined as having never smoked or who reported having smoked less than 100 cigarettes in their lifetime (21). Their eCO had to be < 7 ppm to exclude subjects significantly exposed to cigarette smoke or to environmental sources of carbon monoxide.

Current, former and never smokers had to satisfy the following **inclusion criteria**

- healthy adult subjects (age 18-50 yrs), with no systemic and/or chronic disease
- presence of at least 10 natural anterior teeth (cuspid to cuspid, lower and upper jaw), with no prosthetics or crown

Furthermore, they had to satisfy the following **exclusion criteria**

° Any conditions that could interfere with dental shade measurements, including
  - regular daily use of mouth rinse containing essential oil (EO), cetylpyridinium choride (CPC) or chlorhexidine (CHX) for at least the preceding 7 days before screening visit
  - Subjects wearing fixed or removable orthodontic appliance or prothesis (limited to the 12 natural anterior teeth)
° Significant exposure to passive smoking (excludes current smokers)
° Had undergone dental professional cleaning within 6 months prior to screening
° Pregnancy

### Study Design

This is an observational study with a cross-sectional design to assess dental shade by Vita Easyshade V among three study population (current, former, and never smokers). The study consists of a total of four visit: screening visit, baseline visit at day 0 (Visit 1), short-term follow-up visit at day 7 (±1 days) (Visit 2) and long-term follow-up visit at day 30 (±3 days) (Visit 3) (Figure 1). Subjects will be asked

**Figure 1.**
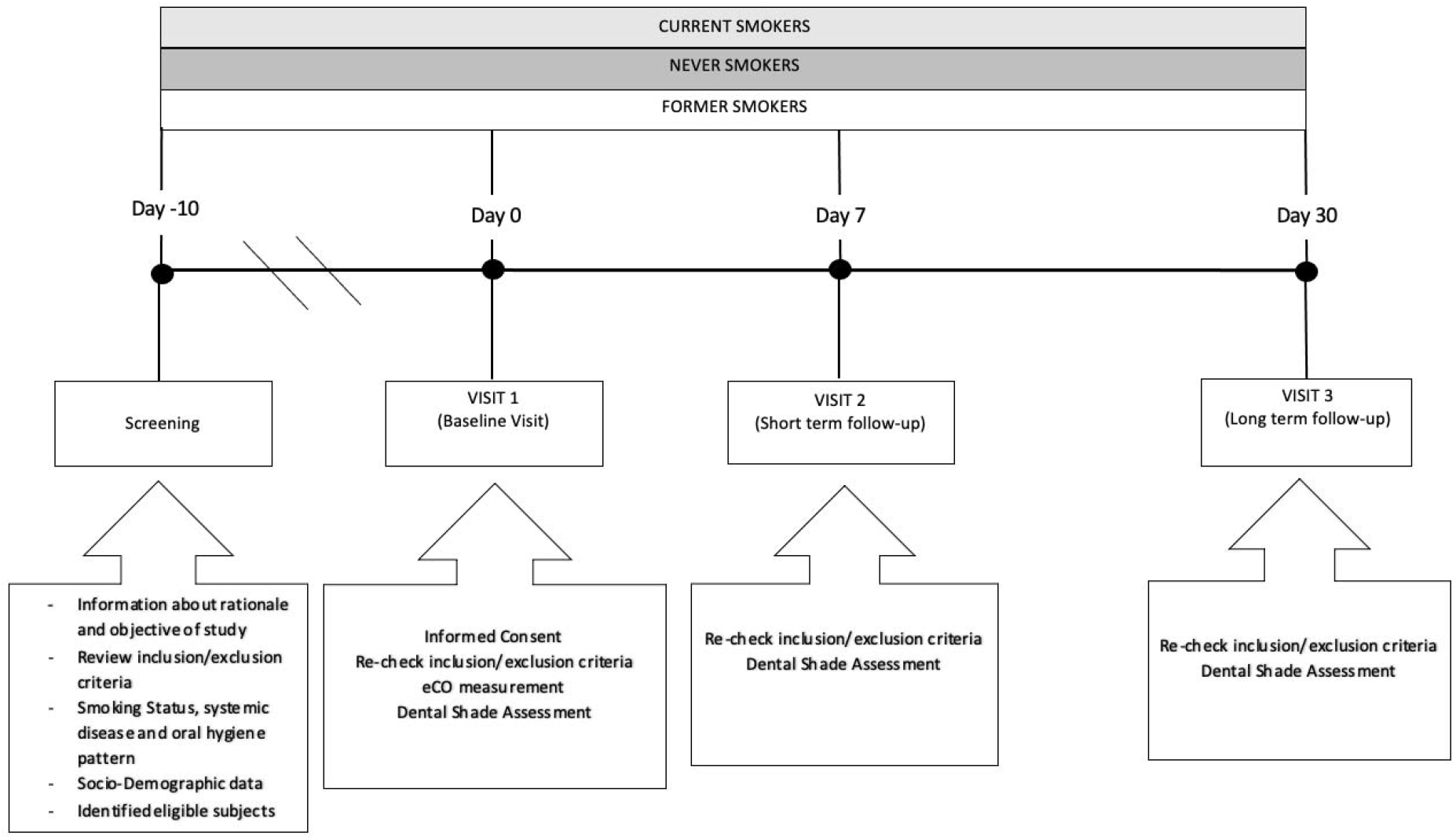
Study Design

- not to change their own habitual oral hygiene (mouthwash, interdental floss) pattern
- for the whole duration of the study
- avoid scaling and polishing procedures for the whole duration of the study
- not to daily use mouthrinse for the whole duration of the study
- not to smoke for at least 2 hrs prior to each study visit
- not to toothbrush for at least 2 hrs prior to each study visit
- not to eat and/or drink (except water) for at least 2 hrs prior to each study visit

### Study Visits

#### Screening Visit

Potential participants will attend a screening visit to 1) receive information about the rationale and objectives of the research; 2) verify eligibility criteria by reviewing their inclusion and exclusion criteria; 3) assess smoking status and oral hygiene habit (i.e. frequency of toothbrushing, type of toothpaste, etc.); and 4) record general socio-demographic characteristics (i.e. sex, age and occupation). All eligible subjects will be invited to participate to Baseline Visit (Visit 1).

## Baseline Visit (Visit 1)

The Baseline Visit will be carried out within 10 days of the Screening Visit. Subjects will be asked to go over a patient information sheet and to sign a consent form. After re-checking inclusion/exclusion criteria and reviewing study restrictions, eCO measurement and dental shade assessment will be carried out and baseline data recorded. Subjects will be instructed not to change their habitual oral hygiene pattern and invited to attend next study visit (Visit 2).

### Day-7 Visit (Visit 2)

Visit 2 will be carried out within 7 (±1) days after Visit 1. Eligibility criteria will be verified again. Dental shade assessment will be repeated for short term repeatability. Subjects will be instructed not to change their habitual oral hygiene pattern and invited to attend next study visit (Visit 3).

### Day-30 Visit (Visit3)

Visit 3 will be carried out within 30 (±3) days after Visit 1. After re-checking eligibility criteria, dental shade assessment will be repeated for long term repeatability.

#### Exhaled Carbon Monoxide Measurement

The smoking status will be objectively verified by measuring exhaled carbon monoxide (eCO) levels (eCO > 7 ppm indicating smoking status) with a portable CO monitor (Micro CO; Micro Medical Ltd, UK). Subjects will be asked not to smoke any cigarettes for at least 2 hours prior to eCO level measurements. Subjects will be invited to exhale slowly into adisposable mouthpiece attached to the eCO monitor as per manufacturer’s recommendations. The value of eCO readings will be noted.

#### Dental Shade Assessment

Prior to dental shade assessment, participants will be asked to rinse their mouth with water and will be subjected to gentle flushing and drying by triple syringe if any food debris still remain on the vestibular aspect of the anterior teeth.

All measurements will be performed in the same examination room by the same operator (GC). According to CIE standard, natural direct daylight will be excluded, and illumination condition will be set at 6500 K and 1000 Lux.

Vita Easyshade V (Figure 2.) will be calibrated according to the manufacturer’s instructions and a sterile mouthpiece will be placed on the optic handpiece for each patient.

**Figure 2.**
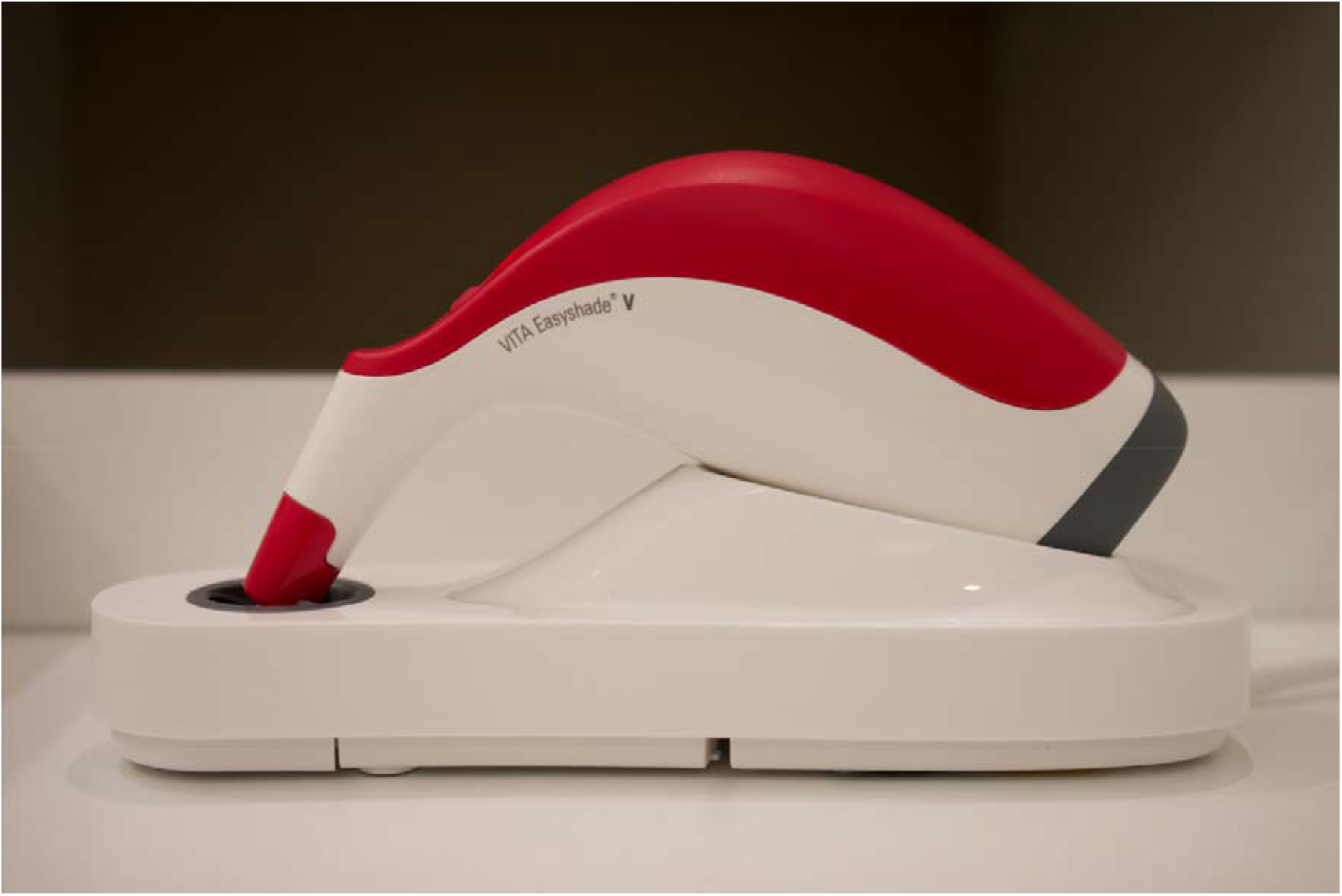
Vita Easyshade V placed on charging station.

The heads of the subjects will be placed against the headrest of the dental chair and their mouths were slightly open, with the tongue away from the anterior teeth.

The shade will be measured at the central tooth area of vestibular surface in “base shade determination” mode with the measuring tip stayed at 90° on the tooth surface to achieve an accurate measurement (Figure 3.). Subjects will be asked to not breathe directly into the measuring tip, causing fogging and inaccurate shade reading.

**Figure 3.**
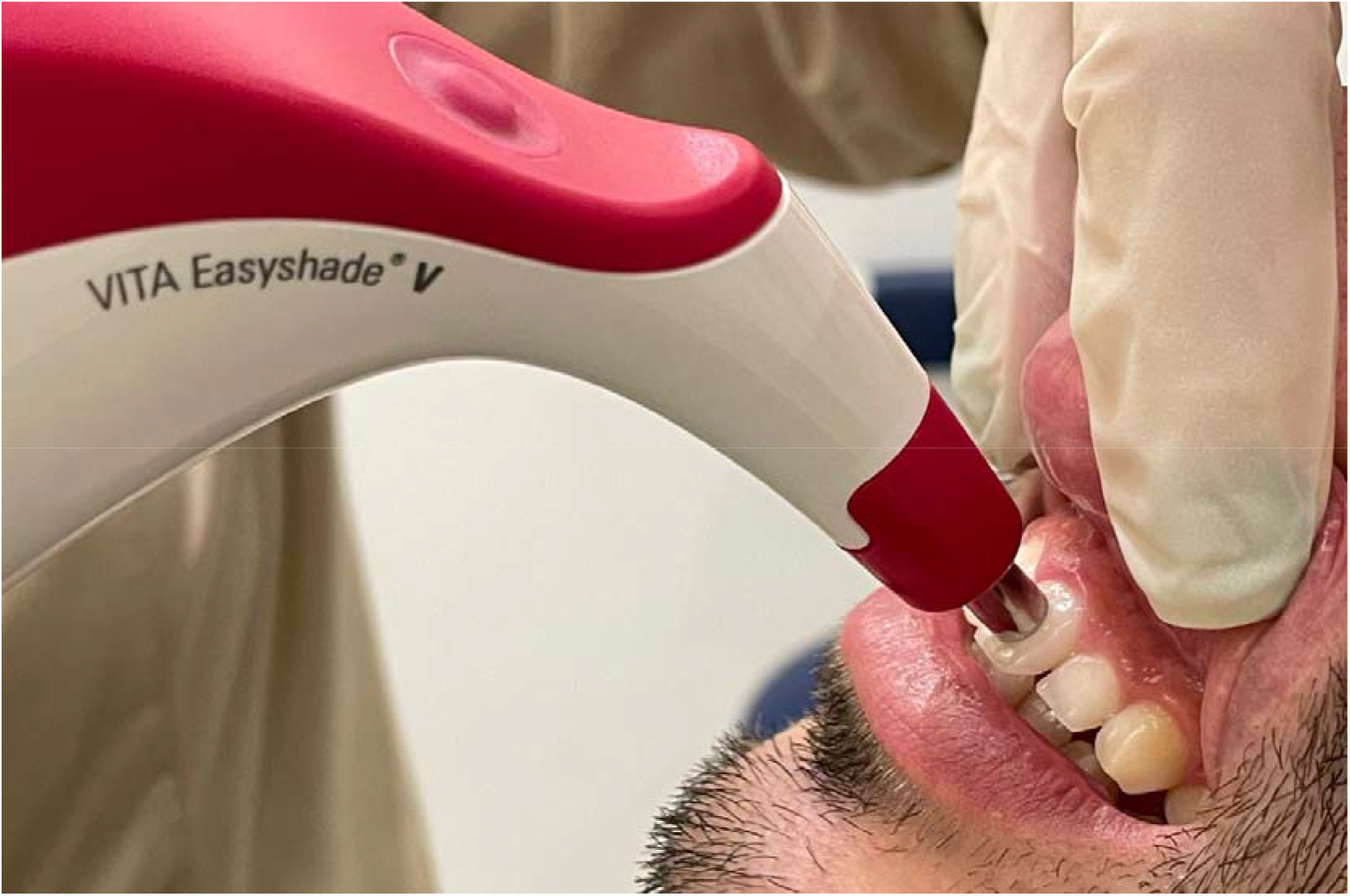
Taking shade measurement in “base shade determination” mode with Vita Easyshade V. The measuring tip stayed at 90° on at the central tooth area of vestibular surface.

CIE L*a*b* color parameters will be calculated and recorded for the vestibular aspect of each anterior tooth (cuspid to cuspid, upper and lower jaw). The coordinate L* is a measure of lightness that ranges from 0 (black) to 100 (white); a* and b* coordinates represent green/red and blue/yellow, respectively.

#### Data Analysis

Short-term repeatability of CIE L*a*b* value will be evaluated by linear regression analysis of measurements obtained at V1 and those obtained at V2 for each study group. Likewise, long-term repeatability will be evaluated by linear regression analysis of measurements obtained at V1 and those obtained at V3. Scatter plots of linear regression analyses will be generated to visualize repeatability results. Moreover, “Bland and Altman” plots will be created to describe the level of agreement between V1 vs V2 and V1 vs V3 for each study groups. 1-tailed sample t test will be also performed to assess the difference from zero of the mean difference between two measurements.

The Range of Normality will be calculated by computing the value corresponding to the mean ± SD*1.64 from the distribution curve of the results of the CIE L*a*b* value in never smokers. Kolmogorov-Smirnov test will be performed to assess the normality data. Categorical data will be summarized by counts and percentages; continuously distributed data, with symmetrical distribution, will be summarized using the mean (standard deviation; SD); continuously distributed data, with skewed distribution, will be summarized using the median (inter-quartile range; IQR). Study groups comparisons will be carried out by Chi-square test, ANOVA and Kruskall-Wallis test for categorical, continuously symmetric and continuously skewed datasets, respectively.

Multiple regression tests will be also performed to identify individual variables, including age, gender, eCO level, pack/years, cig/day, type of toothbrush (i.e. electric toothbrush), type of toothpaste, frequency of toothbrushing, etc. that may influence the results of the dental shade assessment. All analyses will be considered significant with a P-value< 0.05. R version 3.4.3 (2017-11-30) will be utilized for data analysis and generation of graphs.

## RESULTS

Recruitment of participants started in July 2020. Completion of enrolment was achieved in November 2020. Final results will be reported in 1Q 2021.

## DISCUSSION

Avoiding exposure to cigarette smoke toxicants may translate into measurable amelioration of teeth appearance. We will investigate the impact of smoking and smoking cessation on teeth appearance by comparing digital spectrophometry measurements of dental shade in current, former, and never smokers. We will also confirm the validity and reproducibility of dental shade measurements also in these study groups.

In general, dentists rely on shade guides for the visual assessment of natural teeth’s color tone, but shade guides have shown to be unreliable and inaccurate (12,13,14,15). Subjective visual assessment of teeth shades is unsatisfactory for high quality research. More recently, calibrated digital spectrophotometers have been introduced for objective more consistent determination of teeth colors and shades. For this study we will use a popular digital spectrophotometer (VITA Easyshade V). The VITA Easyshade V has been shown to be accurate and reliable in several studies (22,23,24).

By using the VITA Easyshade V digital spectrophotometer (VITA Zahnfabrik, Bad Säckingen, Germany), we intend to measure and compare the CIE L*a*b* values originated by dental shade measurements of maxillary and mandibular cuspids, lateral and central incisors at three different time points in present, former and never smokers.

The issue of test variability is particularly important when investigating dental outcomes in clinical trials of subjects with significant exposure to tobacco smoke (25). To improve measurement reproducibility, we will take great care of the following key confounders by: optimizing of environmental conditions by standardization of ambient light; 2) providing clear instructions about restrictions on all those conditions that could significantly affect study measurements (e.g. variations in oral hygiene practices); 3) having well-trained operators performing the test correctly and accurately; and 4) optimizing CIE L*a*b* color parameters for improved dental shade quantification.

No information is available about changes of CIE L*a*b* values in smokers who quit smoking. Dental discolorations caused by smoking may be permanent, with limited restoration of colors/shades after stopping smoking. If this is true, former smokers should exhibit similar dental shade values as current smokers. The data generated will contribute expanding the existing knowledge base about the impact of smoking on oral health.

Exploring the impact of smoking and smoking cessation on dental aesthetics is important, because for young adult smoker’s bad breath and teeth appearance (due to tooth discoloration and “tar” stains) appears to be much more concerning than the risk of developing cardiopulmonary-related smoking diseases (26,27). For them, this oral health-based narrative may be a much more significant reason to refrain from smoking. That is why we took interest on the quantitation of dental discoloration and objective demonstration of the negative effect of smoking on teeth appearance.

Confirmation of good reproducibility of VITA Easyshade V with clear discrimination of dental shade measurement in current, former, and never smokers will have an important impact on future application of this test for both medical and regulatory research applied not only to combustion-free tobacco products (e.g. e-cigarettes, heated tobacco products, oral tobacco/nicotine products, etc.) and smoking cessation medications, but also to consumer care product for oral hygiene.

